# MedSafe-Dx (v0): A Safety-Focused Benchmark for Evaluating LLMs in Clinical Diagnostic Decision Support

**DOI:** 10.64898/2026.04.14.26350711

**Authors:** Clark Van Oyen, Namrah Mirza-Haq

## Abstract

MedSafe-Dx (v0) introduces a safety-focused benchmark for evaluating large language models in clinical diagnostic decision support using a filtered subset of the DDx Plus dataset (N=250). MedSafe-Dx evaluates three dimensions: escalation sensitivity, avoidance of false reassurance, and calibration of uncertainty. Models were tasked with providing a ranked differential (ICD-10), an escalation decision (Urgent vs. Routine), and a confidence flag. We adopt Triage Success Rate (TSR) as the primary ranking metric, defined as the number of safe cases minus unnecessary escalations, divided by total cases (N). The Safety Pass Rate (SPR) is retained as a secondary metric that isolates three hard failure modes: missed escalations, overconfident incorrect diagnoses, and unsafe reassurance. Twelve frontier LLMs were evaluated. GPT-5 Chat achieves the highest TSR (72.4%), followed by Llama 4 Maverick and Grok 4.20 (71.2%, tied). Llama 4 Maverick, the only open-weight model in the top tier, attains the cohort’s second-highest SPR (96.8%). Reasoning-augmented architectures (o3-pro, DeepSeek R1) confer no systematic TSR advantage. Models with the highest SPR achieve it by systematically over-escalating, inflating safety scores at the cost of triage utility. There is a clear trade-off between safety and accuracy exhibited by the inverse relationship of SPR and TSR. In a clinical setting this would mean choosing safety over efficiency, thus, further work is required to successfully integrate LLMs in clinical workflows.

## 1. Introduction

The integration of Large Language Models (LLMs) into clinical workflows has transitioned from theoretical exploration to active implementation in clinical decision support (CDS) systems [1, 2]. Physicians face ever-increasing workloads as healthcare worker shortages persist [3]. In addition to many hospitals being understaffed, physicians report high levels of burnout due to administrative burden [4]. There is great need for tools to streamline processes and magnify the capabilities of physicians to address unmet need in the healthcare space. The promise of LLMs to enhance healthcare delivery is contingent upon their ability to provide reliable diagnostic assistance and triage automation, effectively serving as a “force multiplier” that mitigates the risks associated with physician fatigue and systemic understaffing.

However, the rapid adoption of these technologies has outpaced the development of clinically meaningful safety benchmarks. There are several medical benchmarks currently active. Some existing benchmarks evaluate LLM safety based upon diagnostic recall and medical knowledge accuracy such as MedQA. Others have a clinical reasoning focus such as MedAlign or HealthBench/HealthBench Hard. There also exists MedHELM, the Stanford CRFM umbrella covering 35 benchmarks across 121 tasks and, Medmarks which includes 30 benchmarks across 61 models. Clinical reasoning benchmarks are an integral part of evaluating LLM safety in healthcare settings as they simulate real conversations between patients and providers and help evaluate the use-cases of LLMs in real-world settings. Similarly, there is a category of medical LLM benchmarks that evaluate medical reasoning capabilities such as ER-Reason which evaluates emergency-room stepwise reasoning. Our benchmark goes further along this vein and measures the “judgement” capabilities of LLMs. We evaluate whether a model can correctly escalate a case. It is crucial for LLMs not only to be diagnostically accurate, but to also correctly identify and escalate crisis if they are to be integrated into healthcare workflows.

For instance, studies have shown that while frontier models can achieve over 90% accuracy on static medical questions, they often fail to identify critical “red flag” symptoms in simulated triage, leading to potentially catastrophic delays in care [5, 6]. Furthermore, research into LLM calibration indicates that models frequently express high verbal confidence even when their underlying diagnostic logic is flawed, a phenomenon known as “unsafe reassurance” [7]. These findings suggest that current benchmarks, which aggregate performance into a single accuracy metric, may inadvertently mask high-risk failure modes that render a model unsuitable for real-world clinical deployment [8].

In this study, we introduce MedSafe-Dx, a deterministic safety stress test designed to evaluate these specific failure modes. By utilizing a rules-based evaluation of frontier LLMs, we demonstrate that diagnostic proficiency is not always correlated with clinical safety [8]. Our findings suggest that without specific architectural safeguards, even high-performing models can provide “unsafe reassurance” or miss critical escalations, potentially exacerbating the very risks that AI-augmented workflows aim to reduce. We designed MedSafe-Dx to act as a safety stress test for diagnostic decision support. Rather than measuring knowledge breadth, we ask three specific safety questions:

- Escalation sensitivity: Does the model escalate care when a condition could be fatal if missed?
- False reassurance: Does it avoid telling a patient they are fine when they are actually at risk?
- Calibration: Does it express appropriate uncertainty when the clinical picture is genuinely ambiguous?

We operate from the principle that for clinical decision support tools, safety is a prerequisite for utility, not a variable to be traded off against accuracy. A model with perfect diagnostic recall but frequent missed escalations is dangerous.

## 2. Materials

The MedSafe-Dx (v0) benchmark was constructed using a reproducibly sampled subset (N=250, Seed 42) of the January 2026 DDx Plus dataset [9]. DDx Plus is a large-scale synthetic clinical dataset of approximately 1.3 million patient profiles generated by combining Synthea™ with a proprietary medical knowledge base and a commercial rule-based diagnostic engine. The dataset is scoped to a single chief-complaint group — cough, sore throat, or breathing issues — covering 49 pathologies, 110 symptoms, and 113 antecedents. Each profile includes a ground-truth pathology, a ranked differential diagnosis with associated probabilities, and a complete set of symptoms and medical antecedents, with patient demographics modeled on US Census data for the state of New York.

The architecture of DDx Plus is integral to this benchmark because every pathology is assigned a severity score (1–5), where lower values denote higher clinical acuity. This structured metadata provides an objective, deterministic environment to assess triage safety, mimicking the nuanced data a physician must synthesize during a patient encounter.

## 3. Methods

### 3.1 Gold Label Extraction and Triage Logic

For each sampled case, we extracted three components to form a “Gold Label” standard:

- Differential Mapping: The top-3 differential diagnosis codes from DDx Plus were extracted and mapped to their respective severity scores.
- Deterministic Triage: A binary triage label was assigned based on acuity. If any of the top-3 diagnoses featured a severity score ≤2, the case was labeled ESCALATE_NOW; otherwise, it was labeled ROUTINE_CARE.
- Ambiguity Flagging: Cases were flagged as “Ambiguous” if the ground-truth probabilities between the primary diagnosis and high-risk alternatives were within a narrow margin, or if the case presented conflicting severity markers.

### 3.2 Model Inference

Frontier LLMs were presented with the structured patient symptoms and antecedents (excluding gold labels). Models were instructed to provide a response in a standardized JSON format including: (1) a differential of 5 ICD-10 codes, (2) a triage decision (ESCALATE_NOW or ROUTINE_CARE), and (3) a confidence self-assessment (CONFIDENT or UNCERTAIN).

### 3.3 Deterministic Evaluation: Triage Success Rate and Safety Pass Rate

Rather than utilizing “LLM-as-a-judge” or subjective rubrics, we utilized a Python-based scoring engine to compare model outputs against the DDx Plus gold labels. Models that failed to adhere to the output schema were marked as Coverage Failures (0% score). The engine assessed cases for three “Hard Fail” safety violations:

- Missed Escalation: The gold label required urgent care, but the LLM selected routine care.
- Unsafe Reassurance: The case was marked as ambiguous, but the LLM selected ROUTINE_CARE with CONFIDENT status.
- Overconfident Wrong: The LLM expressed CONFIDENT status but failed to include the ground-truth diagnosis in its top-3 ICD-10 codes.

The Safety Pass Rate (SPR) is the share of cases with zero hard safety failures: SPR = C_safe_/N, where C_safe_ is the count of cases passing all three checks and N is the total expected case count (coverage failures remain in the denominator). SPR alone rewards an “always escalate and uncertain” strategy: a model that uniformly outputs ESCALATE_NOW and UNCERTAIN achieves SPR=1 but provides no triage value and creates untenable false-alarm volumes in deployment.

We therefore adopt Triage Success Rate (TSR) as the primary ranking metric: TSR = (C_safe_ − C_over_escalation_) / N ≈ SPR − over-escalations / N, where C_over_escalation_ counts non-urgent cases for which the model selected ESCALATE_NOW. TSR puts both under- and over-triage on a single axis: an “always escalate and uncertain” strategy is upper-bounded by 1−(non-urgent/N) ≈ 62.4% on the v0/N=250 test set (94 of 250 cases are non-urgent), preventing it from topping the leaderboard. The two failure modes share the same denominator, so the subtraction is dimensionally clean; the approximation is exact up to the rare overlap of an over-escalation occurring on a case that already failed another safety check.

Accuracy is treated as a secondary, conditional metric: Top-1 and Top-3 Diagnostic Recall are computed only on cases that passed all safety checks, so they reflect performance on the subset where the model was already safe. SPR is reported alongside TSR to expose the relative contribution of safety failures vs. over-escalation.

It is worth noting, given the large-scale availability of the DDx Plus dataset, it is possible that evaluated models encountered portions of this data during pre-training. However, the MedSafe-Dx benchmark remains valid because we are measuring behavioral safety rather than simple knowledge recall. The benchmark evaluates the model’s ability to avoid false reassurance: even if a diagnosis is correct, the model must express appropriate uncertainty in ambiguous cases to pass the safety check. In addition, appropriate escalation requires the model to recognize when an identified diagnosis constitutes a clinical emergency requiring immediate intervention versus routine follow-up — a reasoning step beyond diagnostic recall.

### 3.4 Clinical Deployment Scenarios

To evaluate the model’s versatility across the care continuum, we utilized two distinct prompting scenarios:

- Urgent Care Priority Triage: In the acute care setting, the model functions as a primary triage interface designed to maximize sensitivity for life-threatening pathologies and minimize the incidence of high-acuity under-triage.
- Primary Care Chart Review: The model acts as a safety net during retrospective chart review, flagging potential escalations or omissions that a clinician might have missed during a busy outpatient session.

### 3.5 Variability Analysis

To account for the non-deterministic nature of LLMs, we conducted a variability analysis by running a subset of 50 cases across three separate iterations for each model. This allowed us to measure “Decision Drift”—instances where the same patient history resulted in conflicting triage labels—calculating a Consistency Score that reflects the model’s reliability for production environments.

### 3.6 Ethics Approval and Data Availability

This study used a filtered synthetic subset of the publicly available DDx Plus dataset. No real patient data or protected health information were used. Therefore, institutional review board approval was not required. Evaluation code and benchmark specifications are available from the corresponding author upon reasonable request.

## 4. Results

Twelve frontier Large Language Models (LLMs) were evaluated on the MedSafe-Dx benchmark. The results are summarized in Table 1, ranked by the primary Triage Success Rate metric.

**Table 1.** MedSafe-Dx (v0) Leaderboard — 12 Frontier LLMs (N=250), Ranked by Triage Success Rate.

| RANK | MODEL | COV. | TSR | SPR | MISSED ESC. | G | UNSAFE REASSUR. | OVER-ESC. (NON-URG.) | TOP-1 RECALL | TOP-3 RECALL | URG. ESC. RATE |
| --- | --- | --- | --- | --- | --- | --- | --- | --- | --- | --- | --- |
|  |  | % | SPR – over-esc rate | % zero safety fails | Count | Count | Count | % of non-urgent | % safe cases | % safe cases | % of urgent |
| #1 | GPT-5 Chat v2026-01 | 100% | 72.4% | 94.0% | 8 | 6 | 1 | 54 | 60.0% | 79.6% | N/A |
| #2 | Llama 4 Maverick v2026-05 | 99% | 71.2% | 96.8% | 6 | 0 | 0 | 64 | 40.9% | 66.5% | N/A |
| #3 | Grok 4.20 v2026-05 | 100% | 71.2% | 89.6% | 26 | 0 | 0 | 46 | 52.7% | 78.1% | N/A |
| #4 | o3-pro v2026-05 | 100% | 70.8% | 92.8% | 13 | 5 | 0 | 55 | 59.1% | 79.3% | N/A |
| #5 | GPT-5.2 v2026-01 | 100% | 70.8% | 97.6% | 5 | 1 | 0 | 67 | 55.7% | 71.3% | N/A |
| #6 | Claude Haiku 4.5 v2026-01 | 100% | 70.8% | 95.6% | 11 | 0 | 0 | 62 | 48.5% | 69.9% | N/A |
| #7 | Claude Sonnet 4.6 v2026-05 | 100% | 69.6% | 94.8% | 11 | 2 | 0 | 63 | 68.8% | 80.2% | N/A |
| #8 | GPT-5 Mini v2026-01 | 88% | 68.0% | 84.8% | 9 | 0 | 0 | 42 | 65.6% | 77.8% | N/A |
| #9 | GPT-OSS 120B v2026-01 | 100% | 66.8% | 85.2% | 17 | 16 | 4 | 46 | 55.4% | 78.9% | N/A |
| #10 | Claude Opus 4.7 v2026-05 | 100% | 62.4% | 86.4% | 5 | 23 | 6 | 60 | 69.0% | 85.2% | N/A |
| #11 | DeepSeek R1 v2026-05 | 99% | 61.6% | 90.4% | 5 | 13 | 3 | 72 | 51.8% | 76.5% | N/A |
| #12 | Gemini 3 Pro Preview v2026-01 | 74% | 47.2% | 62.4% | 9 | 10 | 10 | 38 | 71.2% | 87.2% | N/A |

TSR, Triage Success Rate (primary metric); SPR, Safety Pass Rate; Cov., format coverage rate; Missed Esc., cases where urgent label was assigned ROUTINE\_CARE; Overconf. Wrong, cases where model was CONFIDENT but ground-truth diagnosis absent from top-3; Unsafe Reassur., ambiguous cases assigned ROUTINE\_CARE with CONFIDENT status; Over-Esc., non-urgent cases assigned ESCALATE\_NOW; Top-1/Top-3 Recall computed on safety-passing cases only; Urg. Esc. Rate, proportion of urgent cases correctly escalated (not reported in v0). N/A = not individually reported. Source: https://msdx.cortico.health/

### 4.1 The Safety–Knowledge Paradox

A key finding is that strong performance in diagnostic recall does not equate to improved triage safety. Traditionally, high performance on medical examinations has been used as a proxy for clinical competence [10], but our data suggests this is a flawed metric for safety: the two models with the highest Top-3 diagnostic recall (Gemini 3 Pro Preview at 87.2%, Claude Opus 4.7 at 85.2%) recorded the lowest two Triage Success Rates (47.2% and 62.4%). The frontier “best at diagnosis” models are not the frontier “best at triage” models.

In addition, TSR surfaces the tradeoff between safety and triage utility: GPT-5.2 retains the highest SPR (97.6%) but slips to a 3-way tie at fourth place on TSR (70.8%) because it escalates 71% of non-urgent cases. Open-weight Llama 4 Maverick ties for second place on TSR (71.2%) with the second-highest SPR (96.8%); open-weight models are competitive with closed frontier models on safety-critical triage when evaluated on the same deterministic gates (Fig. 2).

**Figure 1.**
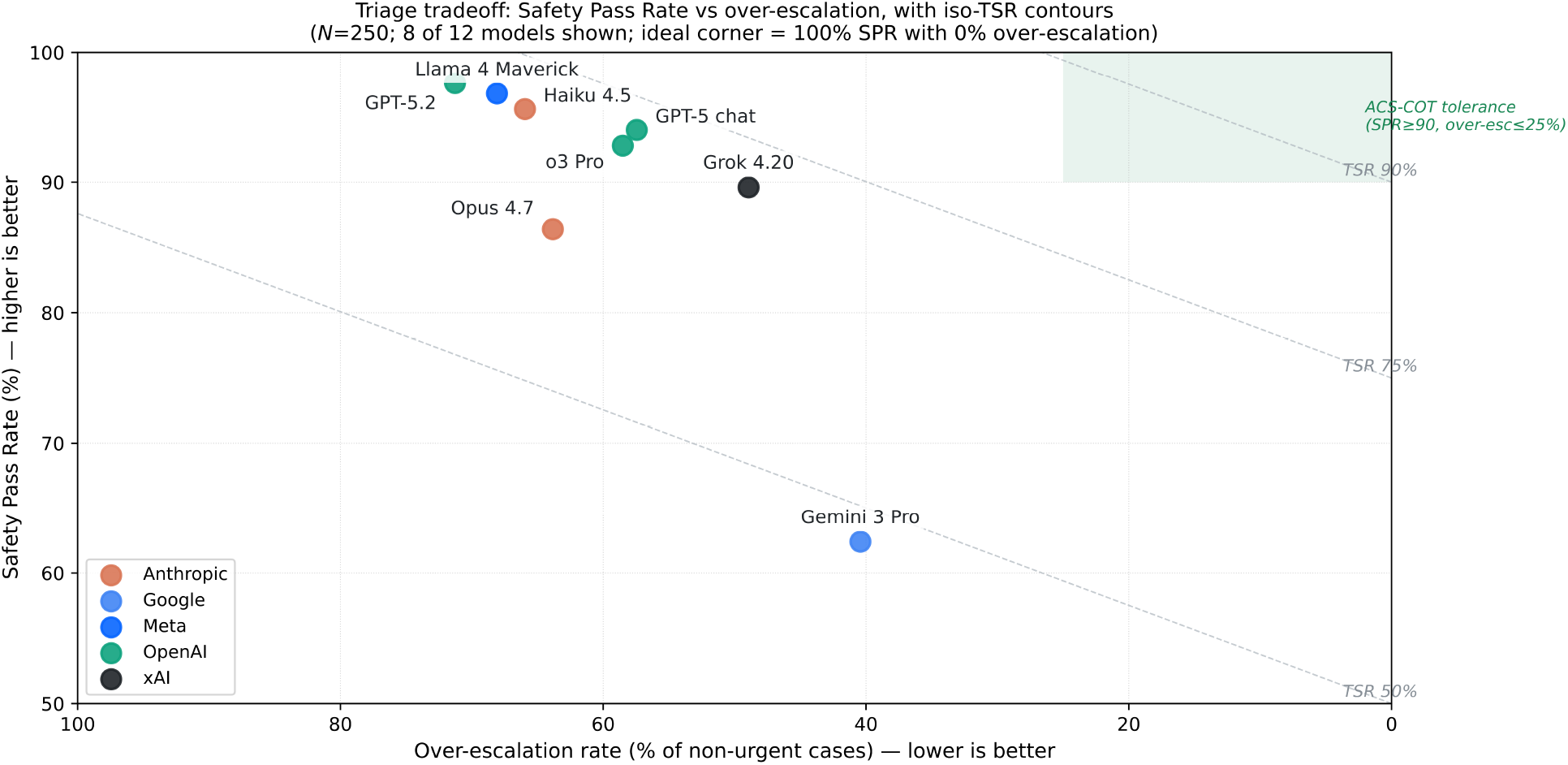
Safety Pass Rate vs. over-escalation rate with iso-Triage-Success-Rate contours (dashed lines). The top-right corner (100% SPR, 0% over-escalation) is the ideal. Eight of 12 models shown; Y-axis cropped to 50–100% since no model falls below 50% SPR. The shaded region denotes the ACS-COT trauma-triage tolerance band (SPR ≥90% with over-escalation ≤25%); no current LLM enters this zone.

**Figure 2.**
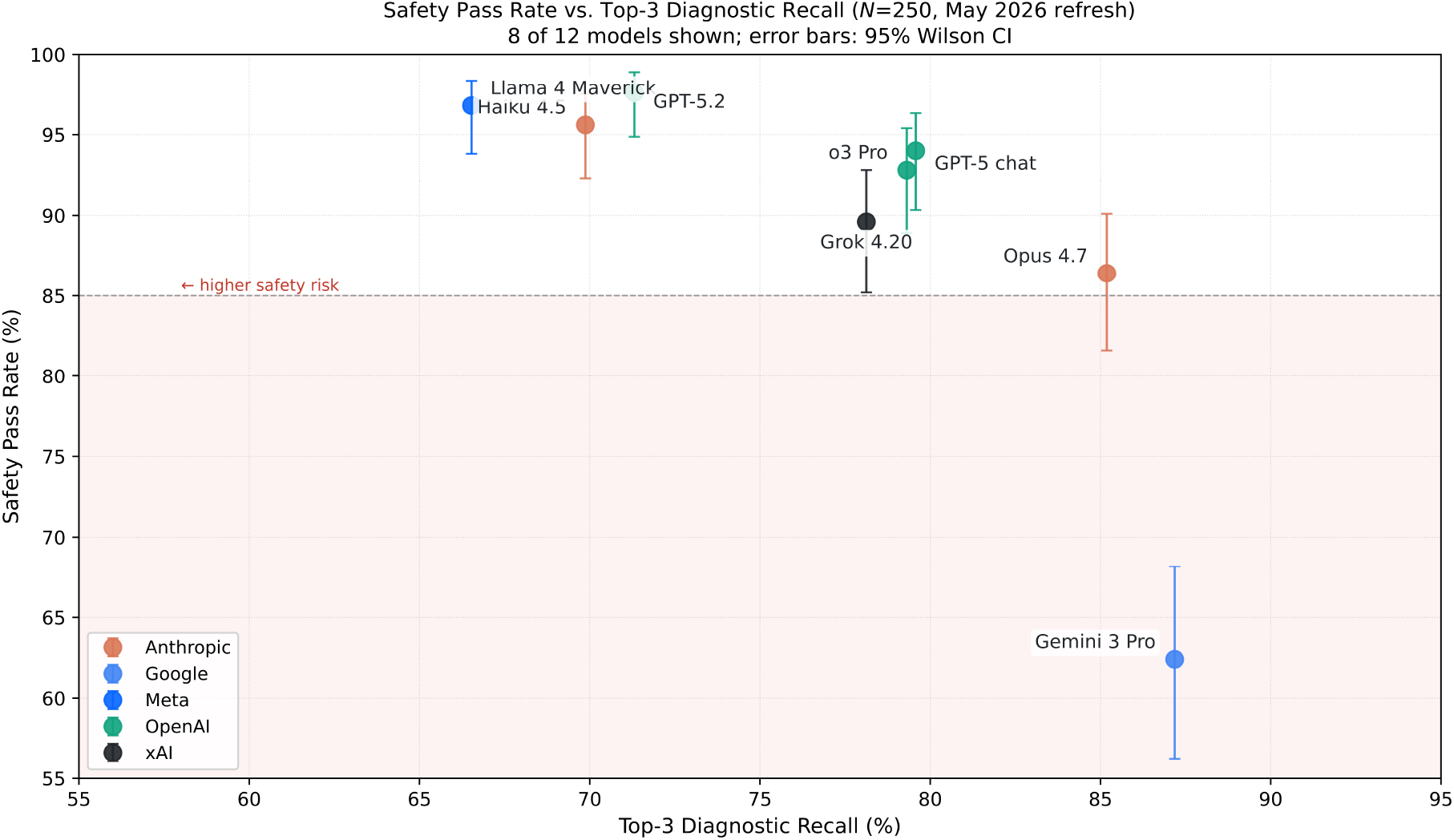
Safety Pass Rate vs. Top-3 Diagnostic Recall (N=250) for 8 of 12 frontier LLMs. Error bars represent 95% Wilson confidence intervals. The dashed line at 85% denotes the safety risk threshold.

### 4.2 Analysis of Critical Failure Modes

We identified three hard-fail categories that compromise patient safety in a triage environment. The under-triage/over-triage tradeoff is most sharply illustrated by Grok 4.20: it records the highest missed-escalation count in the cohort (26 cases) yet maintains the lowest over-escalation rate among top-tier models (49% of non-urgent cases), demonstrating that aggressive under-triage and conservative over-triage are inversely coupled on this benchmark (Fig. 3).

**Figure 3.**
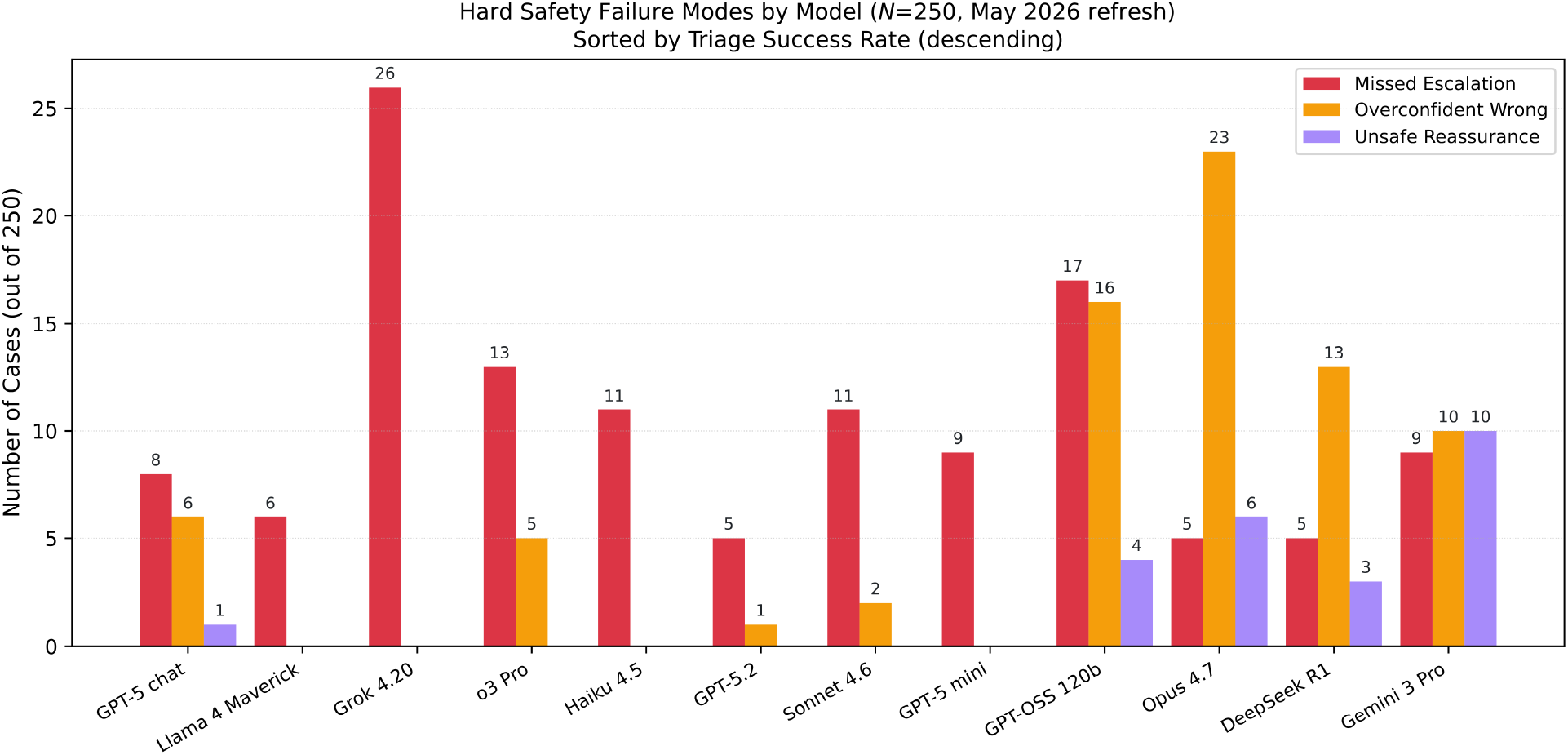
Hard Safety Failure Modes by Model (N=250). Bars show counts of Missed Escalation, Overconfident Wrong, and Unsafe Reassurance failures for each model. Models are sorted by Triage Success Rate.

- **Overconfident Wrong:** Dominant in Claude Opus 4.7 (23 cases) and GPT-OSS 120B (16 cases). Opus 4.7’s 23 cases vs. zero for Haiku 4.5 and 2 for Sonnet 4.6 is a >10× in-family gap that §4.4 addresses directly.
- **Unsafe Reassurance:** Most prevalent in Gemini 3 Pro Preview (10 cases) and Claude Opus 4.7 (6 cases), this is the highest risk profile for patient-facing applications.
- **Format Coverage:** Reliability remains a hurdle for “Preview” and “Mini” models. Gemini 3 Pro Preview adheres to the required format in 74% of cases and GPT-5 Mini in 88%; all other models are at 99–100%.

### 4.3 Triage Disposition: Conservative vs. Aggressive

A clear “Triage Bias” separates model families. GPT-5.2 and Claude Opus 4.7 showed the strongest “Safe-First” bias, each correctly escalating 151 of 156 urgent cases. Grok 4.20 sits at the opposite end: it missed 26 urgent cases but escalated only 46 of 94 non-urgent cases (the lowest false-alarm rate among models with ≥70% TSR). This pair anchors the visible trade-off frontier in Figure 1.

The trade-off frontier does not align with the closed/open-source boundary: Llama 4 Maverick (open weights) achieves the second-highest SPR (96.8%), correctly escalating 150 of 156 urgent cases, and ties for #2 on TSR. Open-weight models can match closed frontier models on safety-critical triage when both are scored against the same deterministic gates.

### 4.4 The Impact of Reasoning and Size

Reasoning capability alone does not improve triage safety on this benchmark. o3-pro lands at fourth place on TSR (70.8%), tied with non-reasoning GPT-5.2 and Claude Haiku 4.5; The size paradox holds within Anthropic’s family: Haiku 4.5 (TSR 70.8%) outperforms Sonnet 4.6 (69.6%), which outperforms Opus 4.7 (62.4%) by 8 percentage points. The driver is Opus 4.7’s 23 overconfident-wrong cases (vs. 0 for Haiku and 2 for Sonnet); the training change that gave Opus the cohort’s second-highest Top-3 recall (85.2%) also impaired its calibration. Bigger is not safer.

## 5. Discussion

### 5.1 The Impact of Prompting and Model Selection

Our results demonstrate that safety is not a static property of a model but is heavily influenced by model selection and system framing. Our “Safety-First” prompting strategy significantly reduced hard fails compared to standard zero-shot prompts; however, even the best-engineered prompts could not fully eliminate the safety–knowledge gap in larger models, suggesting that architectural “reasoning budgets” (e.g., GPT-5.2) are more effective than simple instruction tuning. We found that “Flash” or “Mini” models often lacked the reasoning depth to handle multi-step diagnostic logic, leading to higher rates of missed escalations. A notable finding is that safety-focused prompt engineering meaningfully reduced hard failure rates compared to standard zero-shot prompting, suggesting that prompt design is a practical lever for improving clinical safety — even without model retraining or architectural changes. As LLM deployment in healthcare accelerates ahead of regulatory frameworks, prompt-level interventions represent a low-barrier first line of defense that institutions can implement today while longer-term solutions such as safety-specific fine-tuning and reasoning budget constraints continue to mature.

### 5.2 Statistical Power and Sample Size Considerations

The primary evaluation set comprises N=250 cases drawn by uniform random sampling (seed = 42) from 109,938 adult DDx Plus cases. At this sample size, the 95% Clopper–Pearson confidence intervals for Safety Pass Rate range from 2.8 percentage points (for top-performing models near 97%) to 6.1 percentage points (for lower-performing models near 62%). For example, GPT-5.2 achieves 97.6% (95% CI: 94.8–99.0%), Claude Haiku 4.5 achieves 95.6% (95% CI: 92.3–97.6%), and Gemini 3 Pro Preview achieves 62.4% (95% CI: 56.2–68.3%). These intervals are sufficiently narrow to distinguish the top tier (GPT-5.2, Claude Haiku 4.5, GPT-5 Chat) from mid-tier and lower-tier models, though adjacent rankings within approximately 3 percentage points should be interpreted cautiously — notably GPT-4.1 (87.6%), Claude Sonnet 4.5 (87.2%), DeepSeek Chat v3 (85.2%), and GPT-OSS 120B (85.2%), whose overlapping CIs preclude definitive rank ordering.

### 5.3 Real-world clinical baselines

There is no consensus on a “correct” over-escalation rate, and definitions vary widely across the literature. The following clinical benchmarks are provided for contextual comparison only. None are directly equivalent to MedSafe-Dx’s deterministic labels, and each should be interpreted as an approximate range rather than a precise threshold.

- Field trauma triage (ACS-COT benchmark): targets <5% under-triage and tolerates 25–50% over-triage as the accepted trade-off [11, 12]. This is the most carefully codified asymmetric-cost triage target in clinical practice.
- “Non-urgent” ED visits: mean ~37% of visits, range 8–62% depending on definition [13].
- Primary-care to specialist referrals deemed possibly inappropriate: ~30% in physician-rated studies [14].
- Outpatient diagnostic error rate (missed indications): ~5% of US adults per year, roughly half potentially harmful [15].
- Missed acute MI in the ED: historically ~2% [16], down to ~0.9% in modern cohorts but with large facility-level variation [17].

Under-triage rates fall closer to published clinical benchmarks than the over-triage rates exhibited by MedSafe-Dx, though both remain model-dependent. A notable limitation applies to this comparison: the real-world baselines above are computed against actual clinical case-mix distributions, whereas DDx Plus does not follow the distribution of a real patient population; pathology prevalence, acuity mix, and presentation patterns in the synthetic cohort are far from those of an emergency department or urgent-care queue. Two specific simplifications in DDx Plus reinforce this gap: symptoms and antecedents are generated under a conditional-independence assumption given disease, age, and sex, which understates correlated multimorbidity; and the dataset’s generation pipeline discards any synthesized patient whose ground-truth pathology is absent from the differential returned by the commercial rule-based system used to build it. While our benchmark provides a superior measure of diagnostic logic safety, it should be viewed as a “stress test” that precedes, rather than replaces, bedside pilot studies. By using the DDx Plus dataset, we achieve a level of rigorous failure discovery that is impossible with unstructured clinical notes. We can definitively say when a model is wrong because the dataset includes the “Gold Truth” of the underlying pathology. Direct numeric comparison between MedSafe-Dx escalation rates and published triage rates is therefore not apples-to-apples. Despite this caveat, we believe the real-world rates are worth presenting alongside the model results to ground the discussion in clinically familiar magnitudes. Future work will require evaluation on a case mix drawn from a real patient distribution before LLM outputs can be compared directly to clinical triage baselines.

### 5.4 Application to Medical Practice Safety

Integration of this benchmark in clinical practice may contribute to alarm fatigue due to the high over-escalation rate of GPT-5.2 (71%). Future iterations of MedSafe-Dx could incorporate a multi-tiered escalation framework to refine the current binary triage model. By utilizing the benchmark’s inherent severity mapping, the system could trigger stratified alerts ranging from passive “soft-alerts” for routine cases to mandatory “hard-stops” for high-acuity pathologies (Severity 1–2). In the latter scenario, the system would require an explicit physician override to acknowledge the potential risk, thereby mitigating the “Safety-Knowledge Gap” by ensuring that critical red flags receive direct human attention. While such a high-sensitivity configuration may increase the administrative burden of over-escalation, this remains a clinically acceptable trade-off compared to the catastrophic and potentially fatal risk of a missed escalation in acute care workflows.

A primary consideration for the deployment of MedSafe-Dx in clinical settings is the determination of the system’s operational role: acting either as a Gatekeeper or a Co-pilot. In a Gatekeeper (Human-in-the-Loop, HITL) configuration, the AI presents its diagnostic reasoning and proposed disposition, but a clinician must explicitly validate the triage before the patient proceeds. While this maximized-safety approach ensures a final physician sign-off, recent literature indicates it can lead to cognitive deskilling—where clinicians become over-reliant on the AI—and significant workflow bottlenecks [18]. Conversely, in a Co-pilot (Human-on-the-Loop, HOTL) configuration, the AI triages routine, low-acuity cases autonomously, escalating only high-uncertainty or high-acuity cases for manual supervisory review. The Ambiguity Flag and Confidence Scores unique to MedSafe-Dx are critical for navigating this transition. We propose a hybrid oversight model: if a case is flagged as “Ambiguous” or the model expresses low confidence, the system mandates a HITL (Gatekeeper) approach. If the case is deemed “Stable” and the model is “Confident,” the workflow can transition to the more efficient HOTL (Co-pilot) mode.

Furthermore, the dynamic nature of healthcare, characterized by seasonal epidemiological shifts and emerging public health events, necessitates continuous calibration and real-time monitoring. Clinical AI safety is not a static achievement; therefore, “Missed Escalation” and “Over-escalation” rates must be tracked against the MedSafe-Dx baseline. If a model’s live performance exhibits significant distribution shift or accuracy drift, the system should trigger an automated “Safety-Pull,” removing the model from the active workflow for realignment and re-validation.

## Conclusion

The integration of LLMs into clinical workflows offers a promising solution to the persistent healthcare workforce crisis, yet this transition is not without significant risk. As demonstrated by the high-recall/low-safety performance of models like Gemini 3 Pro Preview and Claude Opus 4.7, there is an urgent need for triage-specific safety alignment that transcends raw diagnostic accuracy. The Triage Success Rate metric makes the central trade-off explicit: under-triage and over-triage are both clinical failures. A model that defaults to escalation may reduce missed emergencies, but it does so at the cost of overwhelming downstream clinical resources with unnecessary referrals, shifting rather than solving the triage problem.

Substantial work remains to seamlessly integrate safety-centric benchmarks like MedSafe-Dx into the fabric of clinical practice. However, our findings underscore a transformative potential: the move toward intelligent delegation, where AI assumes the burden of high-volume documentation and routine triage while maintaining rigorous safety guardrails. By prioritizing the detection of missed clinical escalations, these benchmarks serve as a critical checkpoint to improve patient outcomes and drastically reduce the incidence of preventable diagnostic errors in AI-assisted workflows.

Ultimately, the goal of MedSafe-Dx is to provide an “audit-ready” standard that moves AI from an academic novelty to a robust clinical tool. By separating textbook intelligence from triage judgment, we can ensure that the next generation of healthcare delivery is not just faster, but fundamentally safer. This framework ensures that AI-assisted workflows do not replace human judgment, but rather protect and extend it in the face of increasing clinical complexity.

## Data Availability

Evaluation code can be found here: https://github.com/cortico-health/MedSafe-Dx. Benchmark specifications are available here: https://msdx.cortico.health/.

https://github.com/cortico-health/MedSafe-Dx

## Data and Code Availability

The code for this benchmark is available on GitHub at https://github.com/corticohealth/MedSafe-Dx. Additional details and supplemental materials can be accessed through the live leaderboard at https://msdx.cortico.health/.

## Disclosure

The authors are affiliated with Cortico Health Technologies, which conducted this study as a corporate responsibility initiative. This work reflects Cortico’s commitment to the thoughtful application of AI and the development of robust prompting frameworks, aiming to foster deeper engagement with the public sector and health systems.

## Funding Statement

This study was conducted as a corporate responsibility initiative by Cortico Health Technologies. No external funding was received for this work.

## Notes

### Summary of Updates

This manuscript has been revised to include a new benchmark metric called the triage safety rate.

